# Generating interpretable predictions about antidepressant treatment stability using supervised topic models

**DOI:** 10.1101/2020.03.18.20038232

**Authors:** Michael C. Hughes, Melanie F. Pradier, Andrew Slavin Ross, Thomas H. McCoy, Roy H. Perlis, Finale Doshi-Velez

**Affiliations:** Department of Computer Science, Tufts University, Medford, MA; John A. Paulson School of Engineering and Applied Sciences, Harvard University, Cambridge, MA; Center for Quantitative Health, Massachusetts General Hospital, Boston, MA; Harvard Medical School, Boston, MA

## Abstract

**Importance:** In the absence of readily-assessed and clinically-validated predictors of treatment response, pharmacologic management of major depressive disorder (MDD) often relies on trial and error.

**Objective:** To utilize electronic health records to identify predictors of treatment response, while preserving interpretability of predictions despite large numbers of covariates.

**Design:** Retrospective cohort study.

**Setting:** Two academic medical centers in Boston, including outpatient primary and specialty care clinics.

**Participants:** 81,630 adults with a coded diagnosis of MDD.

**Exposure:** Treatment with 1 or more of 11 standard antidepressants.

**Main Outcomes and Methods:** Stable treatment, intended as a proxy for treatment effectiveness, defined as continued prescription of an antidepressant for 90 days. We trained supervised topic models to extract 10 interpretable covariates from coded clinical data for stability prediction. Then, using data from one hospital system (Site A) we trained generalized linear models and ensembles of decision trees to predict stability outcomes from topic features that summarize patient history. We evaluated on held-out patients from Site A as well as all individuals from a second hospital system (B).

**Results:** Among the 81,630 adults (31% male; age 18-80 with mean 48.46), we identified 55,303 who reached a stable treatment regimen during follow-up. For held-out patients from Site A, mean area-under-the-receiver-operating-characteristic-curve (AUC) discrimination for general stability outcome was 0.627 (95% confidence interval (CI) 0.615 - 0.639) for our supervised topic model with 10 covariates. In evaluation on site B, our approach achieved similar AUC of 0.619 (95% CI 0.610 - 0.627). Building models to predict stability *specific* to a particular drug did not improve upon predicting general stability, even when using a harder-to-interpret ensemble classifier and 9,256 coded covariates (specific AUC = 0.647, 95% CI 0.635-0.658; general AUC = 0.661, 95% CI 0.648-0.672). Topics coherently captured clinical concepts associated with treatment response.

**Conclusions and Relevance:** Coded clinical data available in electronic health records facilitated prediction of general treatment response, but not response to specific medications. While greater discrimination is likely required for clinical application, our results provide a simple and transparent baseline for such studies.

**Funding:** Oracle Labs, Harvard SEAS, and National Institute of Mental Health.

**Links:** Supplement document providing more results, links to interactive visualizations, and detailed procedures for reproducibility https://www.michaelchughes.com/papers/HughesEtAl_medRxiv2020_Supplement.pdf

STROBE checklist: https://www.michaelchughes.com/papers/HughesEtAl_medRxiv2020_STROBE_checklist.pdf

Open-source code for our proposed machine learning methods https://github.com/dtak/prediction-constrained-topic-models

**Key Points:** *Question:* How well can coded clinical data from electronic health records be used to predict achievement of a stable antidepressant regimen in major depressive disorder?

*Findings:* In this in silico cohort study of 81,630 adults, we identified 55,303 who reached a stable antidepressant treatment regimen. Predictions using generalized linear models or ensembles of decision trees applied to diagnosis, procedure, and medication codes, as well as low-dimensional summaries of these codes via supervised topic models, achieved area under receiver operating characteristic curve values of ∼0.62-0.65; treatment-specific models performed no better than general treatment outcome models.

*Meaning:* Coded clinical data can facilitate prediction of antidepressant treatment outcomes, but medication-specific models do not outperform general response prediction models.

## Introduction

Meta-analysis suggests that newer antidepressants are on average very similar in efficacy and overall tolerability^1^a finding further supported by a small number of effectiveness studies^2–4^. However, these group averages obscure a wide amount of interindividual variability; even before recent enthusiasm for precision or personalized medicine, a vast literature addressed potential predictors of antidepressant treatment outcome aimed at identifying individuals who are more or less likely to benefit^5^. For example, symptom-defined subtypes were investigated, initially as predictors of tricyclic antidepressant or monoamine oxidase inhibitor response, then as predictors of selective serotonin reuptake inhibitor response^6–8^. More recently, rather than clinical subtypes, efforts have focused on deriving constellations of symptoms more predictive of response^9–11^or incorporating additional survey measures^12^. Beyond clinical predictors, numerous studies examined incorporation of biomarkers, most notably (and notoriously) the dexamethasone suppression test^13,14^.

A key challenge in all of these studies has been the paucity of head-to-head antidepressant studies, such that it is often difficult to distinguish predictors of poor outcomes overall from predictors of poor outcomes specific to a given medication. Traditional tests of interaction compound this problem, as they are best-powered for opposing effects (i.e., markers associated with better outcome in one group and poorer outcome in another), when in reality this may not comport with biology. Further, even where head-to-head studies exist, there are rarely replication cohorts to follow up initial associations.

In other contexts, electronic health record (EHR) or administrative data sets have been used to predict clinical outcomes, providing sufficiently large real-world cohorts to allow identification and validation of predictors^15–17^. They may offer the further advantage of operating on data already readily available at the point of care, such that clinical adoption does not require the use of new rating scales or measures. In the present study, we sought to apply widely available health record data to understand two questions: first, to what extent can we identify general (i.e. non-specific) predictors of antidepressant response? Second, can we identify treatment-specific predictors, as might be applied to drive a precision medicine approach to antidepressant prescribing?

In so doing, we also investigated a potential solution to a central problem in analysis of large clinical datasets and machine learning for big data more generally: a lack of interpretability^18–20^. While optimized predictions may be useful, the inability to understand what drives these predictions may impede efforts to validate and disseminate them in clinical settings. Moreover, the reliance on individual clinical data points may limit portability if health systems use different procedure or diagnostic codes to reflect the same underlying concepts. Here, we applied a recently-developed supervised topic modeling approach^21^that yields much simpler predictors based on groups of features that retain discrimination and facilitate interpretability.

## Methods

### Study Design

We utilized an in silico cohort drawn from EHR to examine the association between coded EHR available at time of medication prescription for standard antidepressants and subsequent longitudinal outcome of stable treatment with that medication.

The study cohort included individuals with at least one diagnosis of major depressive disorder (MDD, ICD9 codes 296.2x, 296.3x) or depressive disorder not otherwise specified (ICD9 code 311) who received psychiatric care between 1997 and 2017 across the inpatient and outpatient networks of two large academic medical centers in New England (“Site A” and “Site B”). Patients were excluded if age was less than 18 or greater than 80, if the total observation period was less than 90 days, or if there were fewer than 3 total documented visits (of any type, psychiatric or otherwise) in the EHR.

We extracted de-identified patient-level data using the i2b2 server software (i2b2, Boston, MA, USA)^22^. Available patient data included socio-demographic information (age, sex, race/ethnicity), all diagnostic and procedure codes, and all inpatient and outpatient medication prescriptions.

After applying inclusion criteria (Supplemental eFigure 1), a total of 51,048 patients from Site A were included and randomly assigned to training (50%, n=25524), validation (25%, n=12762), and test (25%, n=12762) subsets. 26,176 patients from Site B form an external validation set. The Partners HealthCare institutional review board approved the study protocol, waiving the requirement for informed consent as only de-identified data was utilized and no human subjects contact was required.

**Figure 1:**
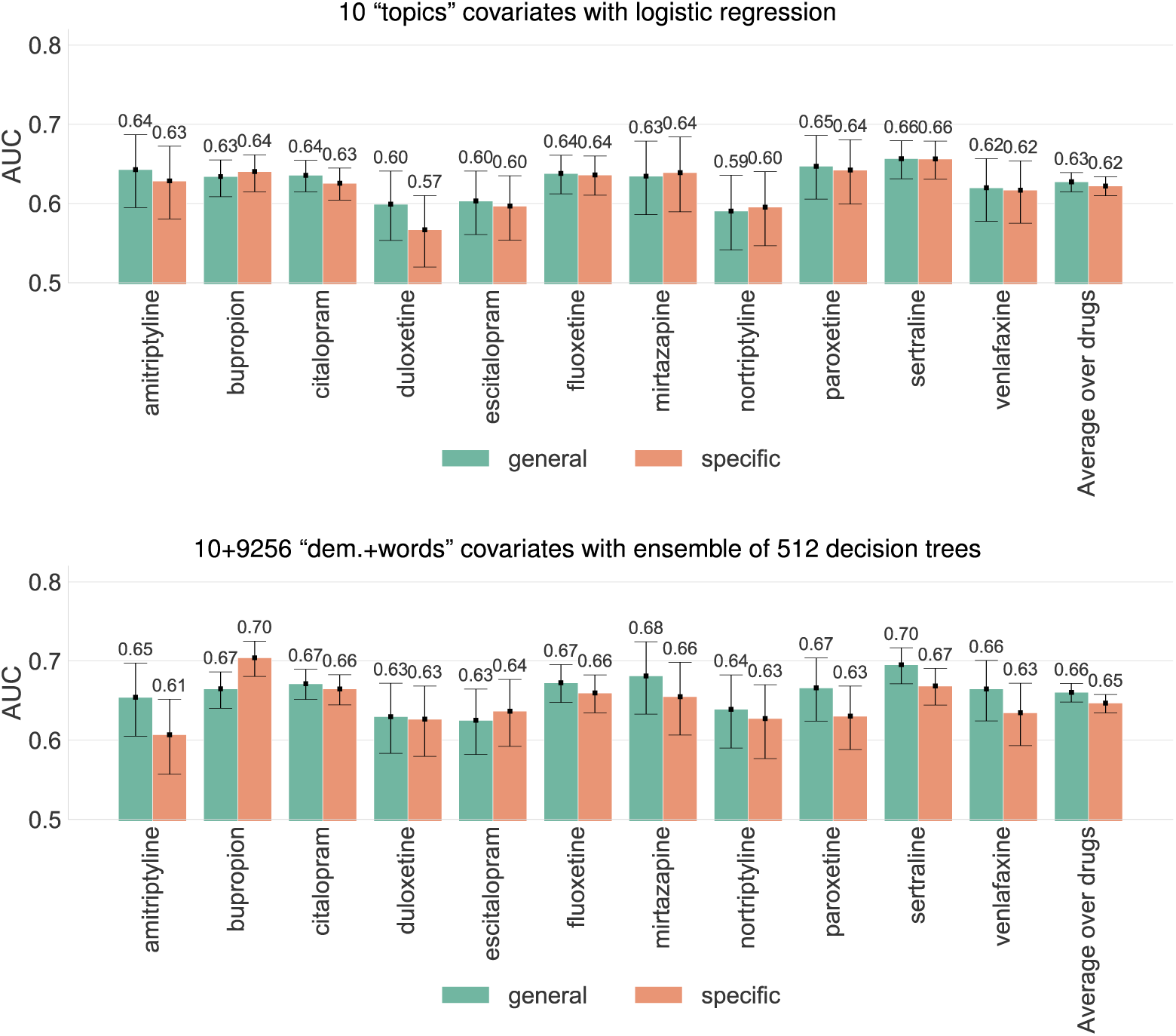
Comparison of General and Drug-specific Stability Prediction for Proposed and Baseline Covariates. Comparison of discriminative ability, as measured by area under the ROC curve (AUC), for general and drug-specific prediction models. Top panel compares models that utilize our proposed 10-dimensional “topics” covariates with a logistic regression predictor. Bottom panel compares models that utilize the baseline high-dimensional “dem+words” covariates with an ensemble of 512 extremely randomized decision trees. For each of the 11 target antidepressants, we obtain an AUC score for a given model by considering predictions from that model on the subset of the Site A test set that includes all known outcomes related to that drug (ignoring data from patients who were never given that drug). To indicate uncertainty in reported AUC values, we repeat our evaluation across 5000 bootstrap samples of each test set and report error bars indicating 95% confidence intervals for the AUC across these bootstrap samples (i.e. the 2.5 and 97.5 percentiles).

### Outcome Definition

Recognizing that traditional clinical trial outcomes such as response and remission are difficult to define reliably for all individuals using solely coded clinical data^16^, we instead sought to identify individuals who achieved a period of stable treatment, as a proxy for ample clinical benefit and tolerability. We applied a simplifying but face-valid assumption that successful treatments continue uninterrupted over time with repeated prescriptions, while unsuccessful treatments are either discontinued or require addition of further medication^4^.

We initially considered 27 possible antidepressants (see Supplemental eTable 1). We defined a treatment segment as stable if it contained at least two prescriptions for the same antidepressant(s) on two distinct dates, the total duration was at least 90 days, the calculated medication possession ratio (fraction of days in segment where the patient possessed a valid, non-expired prescription)^23^was at least 80%, and the largest gap between start dates of any two prescriptions in the segment was between 30 days and 390 days. See Supplemental eFigures 2 and 3 for illustrative examples. Only 11 antidepressants had sufficient usage within Site A (at least 1000 patients) to be used as targets for stability prediction (Supplemental eTable 1).

**Figure 2:**
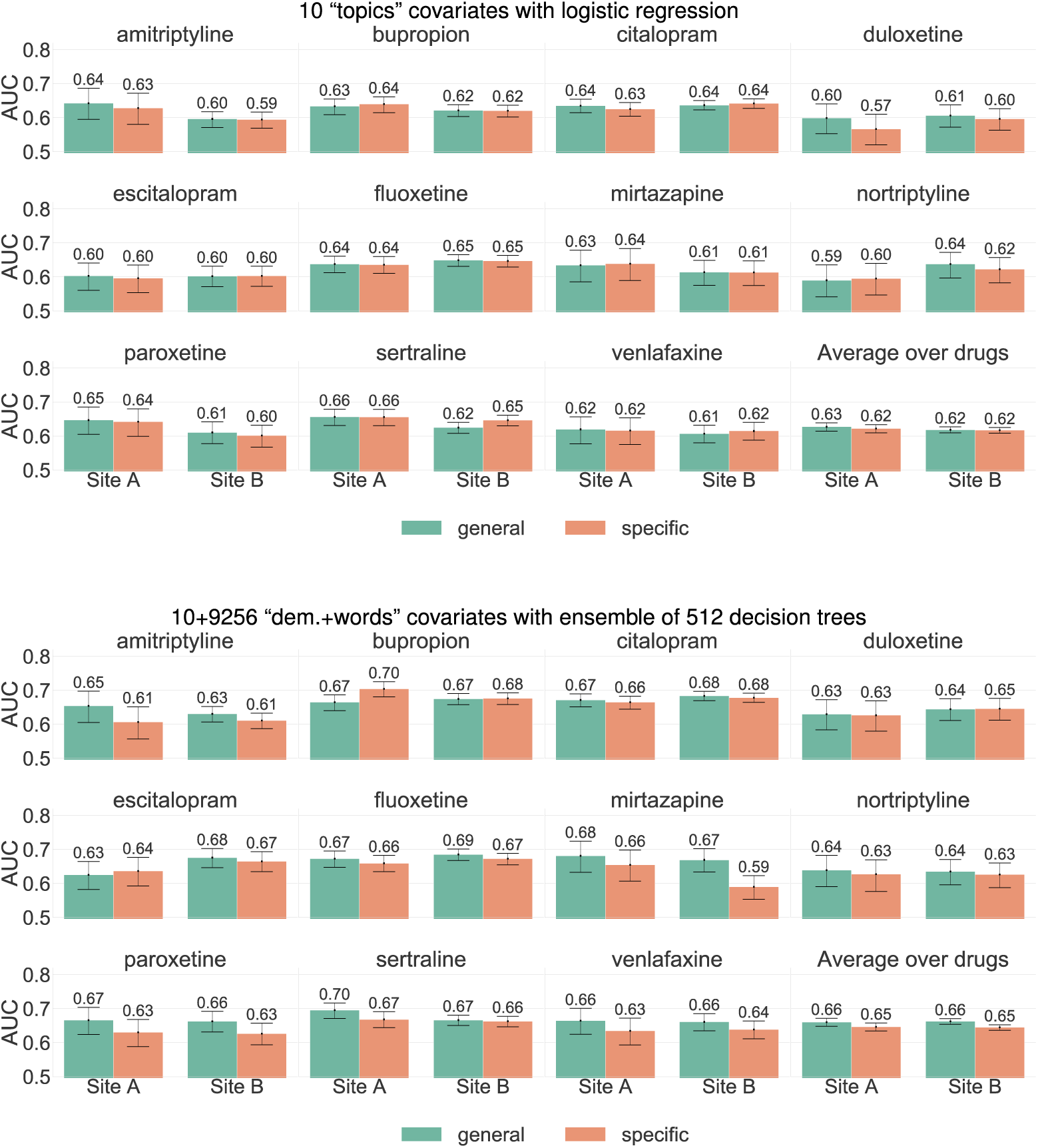
Assessment of Generalization from Site A to Site B for both General and Drug-specific Stability Prediction. Side-by-side comparison of discriminative ability on the Site A and Site B testing sets, as measured by area under the ROC curve (AUC), for general and drug-specific prediction models. Top panel compares models that utilize our proposed 10-dimensional “topics” covariates with a logistic regression predictor. Bottom panel compares models that utilize the baseline high-dimensional “dem.+words” covariates with an ensemble of 512 extremely randomized decision trees. For each of the 11 target antidepressants, we obtain an AUC score for a given model by considering predictions from that model on the subset of the Site A test set that includes all known outcomes related to that drug (ignoring data from patients who were never given that drug). To indicate uncertainty in reported AUC values, we repeat our evaluation across 5000 bootstrap samples of each test set and report error bars indicating 95% confidence intervals for the AUC across these bootstrap samples (i.e. the 2.5 and 97.5 percentiles).

**Figure 3:**
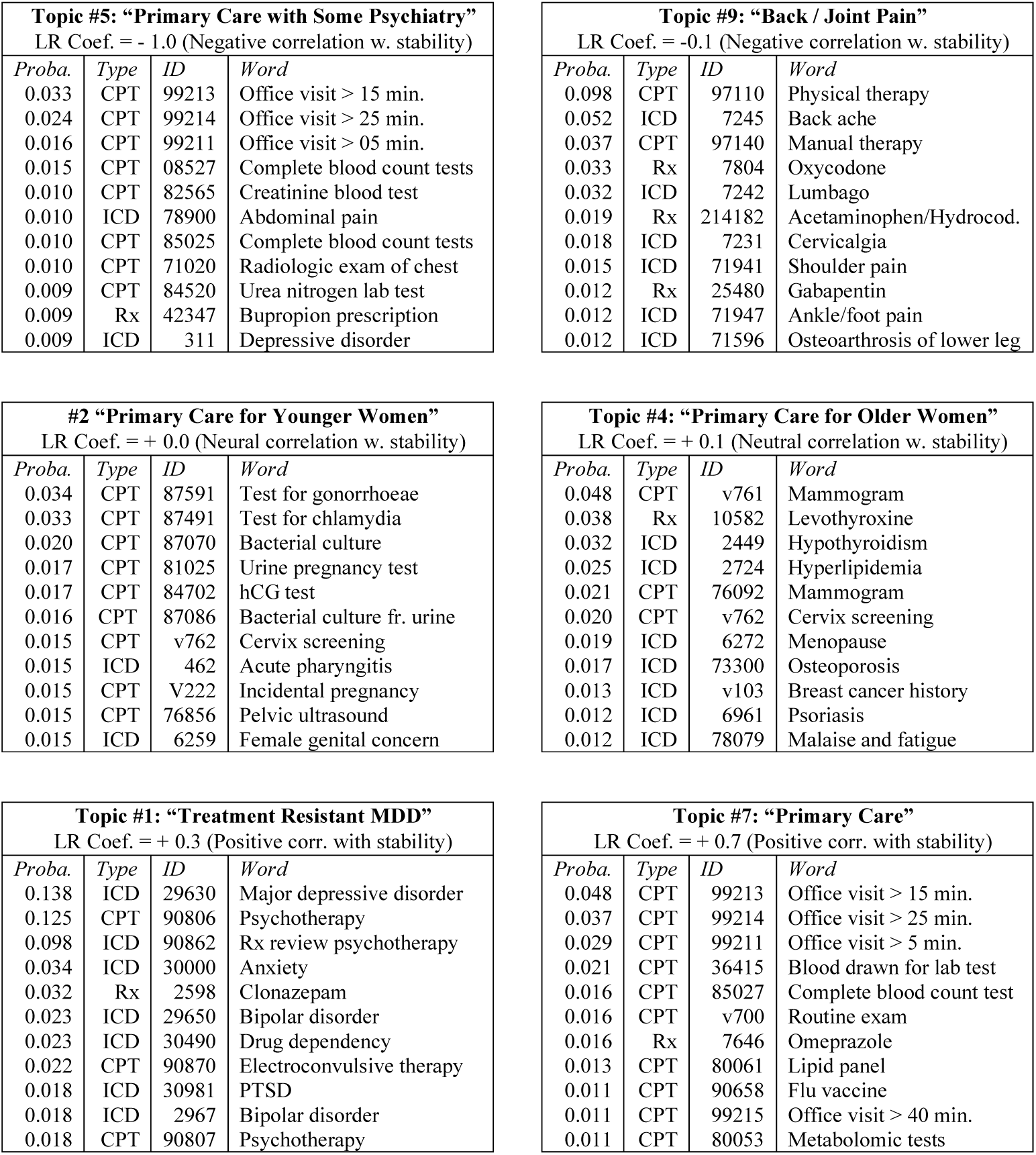
Visualization of Supervised Topic Model for General Stability. Display of 6 learned topics from our proposed PC-sLDA supervised topic model trained to predict general stability. We selected these 6 topics as representative of the 10 total topics learned by the model (top row shows the two most negatively correlated with stability as indicated by each topic’s learned logistic regression coefficient; middle row shows the two most neural; and bottom row shows the two most positively correlated with stability). Each topic is defined by a learned distribution over 9,256 possible diagnostic (ICD), procedural (CPT), and medication-related (Rx) codewords. Codewords with high probability in the same topic are likely to co-occur together in a patient’s record explained by that topic. For each topic, we show its top 10 most probable codes. Each topic is labeled with a clinician annotated title (provided post-hoc by RHP), and the topic’s index order within the original model. We also show the learned logistic regression coefficient (rounded to the nearest 0.1) for the task of predicting general stability. Large positive coefficients suggest that a patient whose history uses more of that topic will be more stable. For an online visualization of all 10 topics, see our [visualization webpage](https://www.eecs.tufts.edu/~mhughes/research/htmlviz/v20190920/rank_words_by=proba_word_given_topic/MODEL_NAME=general-LABEL_NAME=any-N_STATES=10.html).

### Covariate Definition

For each patient, available sociodemographic covariates include sex and race (one-hot categorical) as well as date of the visit and age of the patient (numerical). Additional patient covariates include all available coded billing data (i.e. ICD9 and ICD10 diagnoses, CPT lab tests and procedures), as well as the identity of all prescribed medications. From this initial set of 36,875 possible codes (or “codewords”), we selected 9,256 codewords which occurred in at least 50 patients in Site A. Thus, a count vector of size 9,256 represents a patient’s diagnostic and treatment history.

### Baseline Methods for General and Drug-Specific Stability

The primary aim of prediction was to identify patients likely to exhibit general stability on antidepressants. That is, given the patient’s history up to an evaluation date, predict if the patient will be stable following index prescription of any antidepressant treatment. The secondary aim was to determine whether a subject would exhibit drug-specific stability.

One classifier was trained for the general stability outcome, as well as a separate drug-specific classifier for each of the 11 target antidepressants. We considered two standard probabilistic classifiers, logistic regression and extremely randomized trees, using the open-source implementations in Scikit Learn^24^. All classifiers were trained on Site A’s training set and had hyperparameters selected via grid search on Site A’s validation set to maximize area-under-the-ROC-curve (AUC). Final performance was evaluated on both Site A’s testing set and the independent cohort from Site B.

### Supervised Topic Models for General and Drug-Specific Stability Prediction

A challenge in machine learning is maintaining interpretability while maximizing predictive performance. Even after applying the frequency threshold, an input space of 9,256 codewords limits interpretability and risks model overfitting. We therefore reduced this coded dataset into groups of co-occurring codes indicative of an underlying concept using probabilistic topic models^25^. For a diagram, see Supplemental eFigure 4.

We apply a recent technique for training topic models to perform supervised predictions called *prediction-constrained* topic modeling^21^. Most topic models simply summarize the most salient concepts in the data. For example, diseases such as diabetes, chronic kidney disease, and cancer are prevalent in health records and thus will always be ‘discovered’ as topics. However, it is not clear a priori whether these prominent conditions are relevant to predicting treatment response in MDD - even given the importance of comorbidity, solely rediscovering comorbidity might exclude other features important for prediction. Prediction-constrained (PC) topic models address this issue, finding concepts useful for specific prediction tasks rather than simply summarizing prominent elements. We intend PC topic models to provide low-dimensional patient-specific covariates that yield comparable performance to classifiers that use high-dimensional codeword covariates. However, we hope our topic covariates provide more interpretable insights into how elements of the patient history factor into prediction. For further discussion of topic modeling applications to coded clinical data, see work by McCoy et al.^26,27^.

Following our prior work^21^, we applied prediction-constrained training to fit supervised Latent Dirichlet Allocation topic models (PC-sLDA) to Site A’s training set. We selected 10 topics as representing the best trade-off between validation performance and model size. Experimental details for training and hyperparameter selection, as well as visualization links for trained models, are included in the Supplement. Open-source code is available https://github.com/dtak/prediction-constrained-topic-models.

### Methods for Evaluating Suitability of Models for Medication Prioritization

We further sought to assess how drug-specific models could be used to select medications to prioritize for each patient and compare this to clinical practice. Evaluating such prioritized medications requires certain assumptions, because for most patients we only observed outcomes with one or a few of the 11 possible medications. Given the top 3 suggested medications for a patient, we can assign one of three categories: “unassessable” (none of the 3 have known stability outcomes for that patient), “assessable and stable” (at least one of the top 3 has a positive outcome), and “assessable and non-stable” (none of the top 3 is stable and at least one is non-stable). We can then across a population compute “top-3 stability accuracy” indicating the fraction of assessable patients that would be stable. This evaluation represents a biased but potentially useful proxy for a possible future usage of drug-specific models (biased because models were not trained to prioritize among medications).

### Methods for Evaluating Models as Tools for Forecasting Needed Medication Changes

Finally, we evaluated models of general stability by assessing how well they could forecast the number of medication changes that an individual will require before stability is achieved. For each model, we predict a probability score for each patient in Site A’s test set, use this to stratify subjects into 4 quartiles, then report for each quartile the average number of medication initiations observed in practice before achieving stability.

## Results

Our cohort contained 81630 adults (69% female; age range 18-80 with mean age 48.46) across both sites who met our inclusion criteria based on diagnosis and treatment duration (Supplemental eTable 1). After further filtering out 4,133 patients who lacked any code history before the first visit and thus cannot have personalized predictions as well as 273 subjects from Site B who had no outcomes for the 11 target antidepressants, we had 51,048 patients at Site A (67% female, mean age 48.50) which were further divided into training, validation, and testing sets. Similarly, we had 26,176 patients in Site B (74% female, mean age 48.96) for external evaluation of models. Sociodemographics are summarized in Supplemental eTable 2, with further descriptive statistics in Supplemental eFigure 5.

### Stability Outcome Prevalence and Face Validity

For psychiatrist-treated patients (n=11,985 at Site A), we observed that 2,642 (22%) never reached stability, 5,274 (44%) reached stability on the index prescription, and 4,069 (34%) reached stability eventually. In contrast, for primary care patients (n=14208 at Site A), we observed that 14,208 (34%) were never stable, 19,867 (48%) were stable following index prescription, and 7,583 (18%) were eventually stable. Overall (n=53,643 at Site A), we observed that 16,850 (31%) of patients were never stable, 25,141 (47%) of patients reached stability on the index prescription, and 11,652 (22%) reached stability eventually.

### Comparison of Feature Representations on Site A

Figure 1 compares general and drug-specific prediction models for two possible feature representations: high-dimensional codeword count vectors plus demographics (“dem+words”) and the low-dimensional “topics” covariates provided by our PC-sLDA model. General stability performance was best with “dem+words” features and an ensemble of 512 decision trees, achieving an average AUC of 0.661 (95% CI 0.648 - 0.672). When using a simpler logistic regression classifier, the high-dimensional “dem+words” features yield an average AUC of 0.628 (95% CI 0.614 - 0.639). The 10-covariate topic representation captures much of this discriminative capability even when using simple logistic regression, achieving average AUC 0.627 (95% 0.615 - 0.639). See Supplemental eFigure 6 and eTables 3, 4, 5, and 6 for thorough comparisons of all feature-classifier combinations on Site A.

### Generalization of Stability Outcome Predictions to Site B

Next, we examined the transferability of models trained on data from Site A to separate patients from Site B (see Supplemental eTable 2 for sociodemographics). Distribution of stability outcomes for Site B was similar: across all 27,987 subjects 13,018 (47%) were stable on the index prescription, 5,492 (20%) were stable eventually, and 9,477 (34%) were never stable, compared to 47%, 22%, and 31% at Site A.

Figure 2 shows general stability prediction for both Site A and Site B, again comparing high-dimensional “dem+words” features to our 10-dimensional topic features. Models trained on Site A transfer to Site B with only modest decay in AUC for both feature representations. Using “dem+words” features, average AUC was 0.661 (0.648 - 0.672) for Site A and 0.663 (0.654 - 0.671) for Site B. Using our 10-dimensional topic features, average AUC was 0.627 (0.615 - 0.639) for Site A and 0.619 (0.610 -0.627) for Site B. As an alternative evaluation, Supplemental eFigure 7 plots positive predictive value versus negative predictive value for each model and site.

### Model Interpretability Qualitative Evaluation

We sought to understand which features were important for stability prediction. Figure 3 visualizes representative topics learned by our proposed 10-topic model for general stability. All topics demonstrated sufficient coherence to enable a qualitative description annotated by a clinician (RHP). For example, while topics 5 and 7 both capture routine primary care visits, topic 5 reflects more terms related to psychiatric evaluation, suggesting more aggressive intervention or more severe illness. Topic 1 includes terms indicative of treatment resistance. Topics 2 and 4 capture gynecologic outpatient practice and menopause, respectively. Supplemental materials include hyperlinks to an online visualization tool to explore the important features of all trained models; Supplemental eFigure 8 shows important features for the “dem+words” classifiers.

### Evaluation of Medication Prioritization and Comparison with Clinical Practice

Next, we evaluated the top-3 stability accuracy achieved by models used to prioritize antidepressants for a patient (Supplemental eTable 7). When always predicting the same 3 medications most commonly stable in Site’s A training set, we measured top-3 stability accuracy at 0.602 (95% CI: 0.591-0.612; 64.1% of the 12762 patients in Site A’s test set were assessable). For observed clinical practice (in which many treatments prescribe just one medication, but some prescribe more), the top-3 stability accuracy was 0.602 (95% CI: 0.593-0.611; 99.5% of 12762 patients assessable). This improved to 0.637 (95% CI: 0.628-0.646; 99.8% of 12762 patients assessable) if we allow prescriptions with less than 3 medications to be filled up to 3 total by selecting from the most commonly stable antidepressants. By comparison, our extremely randomized trees model using all demographic and diagnostic code features achieved a top-3 accuracy of 0.622 (95% CI: 0.610-0.634; 47.4% of 12762 patients assessable). Performance with our topic model was poorer: top-3 accuracy was 0.581 (95% CI: 0.566-0.594; 38.2% of 12762 patients assessable).

### Evaluation of General Stability Prediction to Forecast Needed Medication Changes

Finally, we assigned all individuals in the test set to a stability risk quartile by their general stability probability score (Supplemental eTable 8). For our extremely randomized tree model using all demographics and codewords, those in the top quartile had a mean number of additional medication trials of 0.736 (95% CI 0.688-0.796) beyond the initial prescription at first visit to achieve stability. Those in the bottom quartile require an average of 1.754 medication trials (95% CI 1.681-1.843) beyond the initial prescription to achieve stability. By comparison, using our topic model features and LR classifier, the top quartile has a mean number of additional medication trials of 0.864 (95% CI: 0.816-0.918), while the bottom quartile has a mean of 1.722 (95% CI: 1.647-1.799).

## Discussion

In this analysis of electronic health records from more than 81,000 individuals across two health systems, we identified machine learning models that predict achievement of treatment stability, a proxy for effectiveness, based solely on coded clinical data already available rather than incorporating research measures or questionnaires.

At first blush, the discrimination we report is modest, with AUC’s in the 0.60 - 0.66 range. A key question to ask, however, is ‘compared to what?’ We were unable to identify any similar published studies in generalizable cohorts, so we cannot make a direct comparison to another method. While an AUC of 0.8 is often seen as a magic number, we and others have argued that this makes little sense, because the necessary discrimination depends critically on the context in which prediction will be applied^28,29^.

We also found that, contrary to our hypothesis, development of treatment-specific predictors rather than general predictors did not meaningfully improve prediction. This may reflect the observation that much of antidepressant response may be considered placebo-like, or nonspecific. That is, while antidepressants consistently demonstrate superiority to placebo^1^, placebo response is substantial such that nonspecific predictors may outperform drug-specific ones. This result is consistent with the lack of success of efforts to find treatment-specific pharmacogenomic predictors^30^. Our results do not preclude the existence of such medication-specific predictors but suggest that other strategies may be required to identify them.

We also present a framework for understanding the behavior of our drug-specific models if used to guide antidepressant selection, comparing performance to observed clinical practice and to a baseline in which all patients receive the most common antidepressants. It bears emphasis that this represents an instance of transfer learning: the models were not trained to recommend antidepressants per se, nor to mimic clinician performance. However, it illustrates a likely application of these models in practice to personalize treatment selection. We find that the difference between clinician performance and suggesting the one-size-fits-all most common medications is remarkably modest - on the order of 3%. In light of the known similarities in efficacy between standard treatments, essentially all of which are derived from a common set of assumptions about monoaminergic neurotransmission, this is not surprising. Despite enthusiasm about personalized medicine, the hypothesis that personalization improves outcome has rarely been rigorously tested. However, the observation that our best models already yield results similar to clinicians suggests that clinical performance may not be as out of reach as AUCs alone might indicate.

Our analysis also suggests that general stability prediction may still be useful for stratifying patients and understanding personalized chances of stability. We describe an approach to estimating the number of treatment trials that may be avoided, or saved, where models are applied. The top quartile of predicted stability requires about 1 less medication trial than the bottom quartile, which suggests that devoting more care resources - for example, more intensivecare management or scalable evidence-based therapies - to those in the lower quartiles might be a worthy targeted investment.

Our results further indicate that, while topic modeling does not improve prediction over high-dimensional representations, it does yield readily-interpretable concepts relevant to prediction. EHR data is widely acknowledged to be noisy, with codes applied inconsistently even by individual clinicians; it is easy to learn predictors that are capturing a site effect or that simply serve as proxies for some other variable. (Conversely, the individual coded terms are inconsistent between linear and nonlinear models, and many are difficult to align with clinical practice, further illustrating the advantage in interpretability of topic-based models.) The approach we describe, which maps EHR dimensions into interpretable topics, allows stakeholders to easily inspect the learned topic features to understand what co-occurring codeword features in patient history drive predictions. This property is critical for researchers seeking to understand more complex models, but also ultimately for clinicians who may employ them: nominating treatments without understanding why they are favored is unlikely to be accepted by clinicians accustomed to their own strain of personalization^31^. The transferability of our results to a second health system suggests a further advantage, namely that topics may be more robust to overfitting than individual token-based approaches. In other words, if the goal is to build models that generalize across health systems, supervised topics may help to avoid the tendency of code-based models to fit site-specific use of individual procedure or diagnostic codes.

We emphasize multiple important limitations in interpreting these results. The outcome we examined, stability, is markedly different from standard outcomes in clinical trials like remission or 50% reduction in symptoms. The standard approach to using EHR data has attempted to impose a clinical trial-like structure and outcome measure – that is, to extract or impute measures of depression severity. Some studies have sought to emphasize a common primary care depression screening tool such as the PHQ-9^32^, which characterizes symptom frequency, not severity, and was not designed to measure response. Others rely on text from narrative clinical notes^16^. However, these approaches minimize the strengths (availability of large scale, if imperfect, data that corresponds to real-world experience) while emphasizing the weaknesses (lack of precision in diagnosis and symptom measurement) in health records. Moreover, they perpetuate the myth that depression symptoms are purely episodic; in reality, such symptoms tend to wax and wane over time for many patients.

In contrast to these past efforts, we employed a simple metric to assess stability based on historical prescribing data, relying on the fact that clinician and patient behavior is likely to be reasonable: effective and well-tolerated treatments are continued, while ineffective or intolerable medications are discontinued. We are attempting to answer a simple question: “If I write a prescription today, how likely am I to continue writing it for the next 90 days?”

In sum, these results should be considered a starting point: incorporation of additional outcomes, as well as additional clinician- and patient-level predictors, should only improve the quality of prediction. Indeed, improving prediction of individual treatment response will almost certainly require data from multiple modalities. If such predictors are integrated with coded data to form topics, it should be possible not only to achieve greater discrimination while preserving portability, but also to understand the key features driving that discrimination in a way not possible with other machine learning strategies. Once such models emerge, prospective investigation will be needed to determine to what extent they meaningfully improve outcomes, if at all. The present study is a necessary first step towards such investigation.

## Data Availability

Study data consisted of deidentified electronic health records from academic medical centers, but cannot be made available in general due to IRB restrictions. Please contact the authors with questions or concerns.

## Acknowledgements

This study was funded by Oracle Labs and the National Institute of Mental Health (1R01MH106577-01) (R56MH115187). We thank Victor Castro for preparing the deidentified EHR dataset and Partners Research Computing for computational resources. We thank the Center for Research on Computation and Society (CRCS) and Harvard Data Science Initiative (HDSI) for sponsoring Melanie F. Pradier.

